# Seroprevalence of SARS CoV-2 antibodies in healthcare workers and administration employees: a prospective surveillance study at a 1,400-bed university hospital in Germany

**DOI:** 10.1101/2020.09.29.20203737

**Authors:** Christina Bahrs, Aurelia Kimmig, Sebastian Weis, Juliane Ankert, Stefan Hagel, Jens Maschmann, Andreas Stallmach, Andrea Steiner, Michael Bauer, Wilhelm Behringer, Michael Baier, Miriam Kesselmeier, Cora Richert, Florian Zepf, Martin Walter, André Scherag, Michael Kiehntopf, Bettina Löffler, Mathias W. Pletz

## Abstract

**Background:** Healthcare workers (HCWs) are at particular risk to acquire SARS-CoV-2 infections.

**Aim:** The objectives of this study were to compare SARS-CoV-2 IgG seroprevalence and compliance to wear personal protective equipment (PPE) between HCWs working within (high-risk) or outside (intermediate-risk) units treating suspected or confirmed COVID-19 patients. In addition, administration staff (low-risk) was included.

**Materials:** Co-HCW is a prospective cohort study among employees at the Jena University Hospital, Germany. Since 16^th^ March 2020, 50 SARS-CoV-2 inpatients and 73 outpatients were treated in our hospital. Mandatory masking was implemented on 20^th^ March 2020. We here evaluated seroprevalence using two IgG detecting immunoassays, assessed COVID-19 exposure, clinical symptoms and compliance to wear PPE.

**Findings:** Between 19^th^ May and 19^th^ June 2020 we analysed 660 employees [out of 3,228; 20.4%]. Eighteen participants (2.7%) had SARS-CoV-2 antibodies in at least one immunoassay. Among them, 13 (72.2%) were not aware of direct COVID-19 exposure and 9 (50.0%) did not report any clinical symptoms. We observed no evidence for an association between seroprevalence and risk area (high-risk: 2 of 137 HCWs (1.5%), intermediate-risk: 10 of 343 HCWs (2.9%), low-risk: 6 of 180 administration employees (3.3%); p=0.574). Reported compliance to wear PPE differed (p<0.001) between working in high-risk (98.3%) and in intermediate-risk areas (69.8%).

**Conclusion:** No evidence for higher seroprevalence against SARS-CoV-2 in HCWs working in high-risk COVID-19 areas could be observed, probably due to high compliance to wear PPE. Compared to administration employees, we observed no additional risk to acquire SARS-CoV-2 infections by patient care.

## Introduction

Severe acute respiratory syndrome coronavirus 2 (SARS-CoV-2) is a novel beta coronavirus that was first identified in December 2019 in Wuhan, China [1, 2]. As of the beginning of 2020 the outbreak progressed and has been characterized as a pandemic in March 2020 [3, 4]. The clinical presentation of the disease caused by SARS-CoV-2, corona virus disease 2019 (COVID-19) [4], varies significantly and ranges from asymptomatic and mild to critical courses [5–7]. As asymptomatic or pre-symptomatic patients can spread the virus [8–11], it is challenging to timely identify and isolate respective cases. Due to the pandemic the disease and health care burden remains high in many parts of the world (https://www.ecdc.europa.eu/en/geographical-distribution-2019-ncov-cases). In Summer 2020, there were more than 1.3 Mio healthcare workers (HCWs) who have been tested positive for SARS-CoV-2 worldwide [12].

SARS-CoV-2 is highly transmissible from human to human mainly via inhalation of infectious respiratory droplets but also via close personal contact (shaking hands) and via touching contaminated surfaces [2, 13]. As a consequence, nosocomial transmission of insufficiently protected HCWs can occur during aerosol generating procedures [13–14], in the regular patient contact particularly when exposed to patients with a delayed diagnosis of COVID-19 and also in close contact with asymptomatic but virus carrying colleagues [15–19]. Hence, it is of great importance to implement infection prevention programs and provide HCWs with sufficient personal protective equipment (PPE) in order to reduce nosocomial transmissions [20]. Further reason for HCWs to be tested positive for COVID-19 is community transmission in particular due to local outbreaks [21], and infectious returning travelers [2]. On 11^th^ March 2020, the first COVID-19 cases were detected in the city of Jena, Germany. Only five days later, a HCW returning from skiing in Austria caused the first nosocomial outbreak at the Jena University Hospital (JUH) that involved further 31 newly confirmed SARS CoV-2 infections until 19^th^ March.

The primary objective of this study was to investigate the spread of SARS-CoV-2 infection by testing for SARS CoV-2 IgG antibodies among employees of JUH with and without patient contact. Secondary objectives were to compare seroprevalence rates between employees working at different COVID-19 risk areas according to working place and to provide insight into the effectiveness of and compliance with the use of PPE.

## Methods

### Study design and setting

The Co-HCW study (SARS-CoV-2 seroprevalence and infection status in employees at JUH) is a prospective, single centre observational cohort study conducted at JUH, a 1,400-bed academic hospital in Germany. Since 16^th^ March 2020, 50 SARS-CoV-2 positive patients were hospitalized and additional 73 SARS-CoV-2 positive outpatients were seen at the emergency department (valid at 8^th^ June 2020). Due to the increasing number of SARS-CoV-2 cases (in Jena and at JUH), mandatory masking for all employees at JUH was implemented on 20^th^ March 2020. Research was conducted in accordance with the declaration of Helsinki and national and institutional standards. The study protocol was approved by the local ethics committee of the Friedrich-Schiller-University Jena (approval no. 2020-1774) and the study was registered at the German Clinical Trials Register (DRKS00022432).

### Enrolment and data management

Participants were recruited between 19^th^ May 2020 and 19^th^ June 2020. All employees of JUH working at predefined areas with (HCWs) and without patient contact (administration employees) (as provided in ***Supplementary Table 1***) were eligible for inclusion. Eligible staff members received information on the planned study per e-mail and/or were directly informed per an on-site visit. Participation was voluntary. Employees were included if they signed a written informed consent form, answered a questionnaire (see below) and agreed on providing a blood sample (not exceeding 9 ml of venous blood).

**Table 1.**
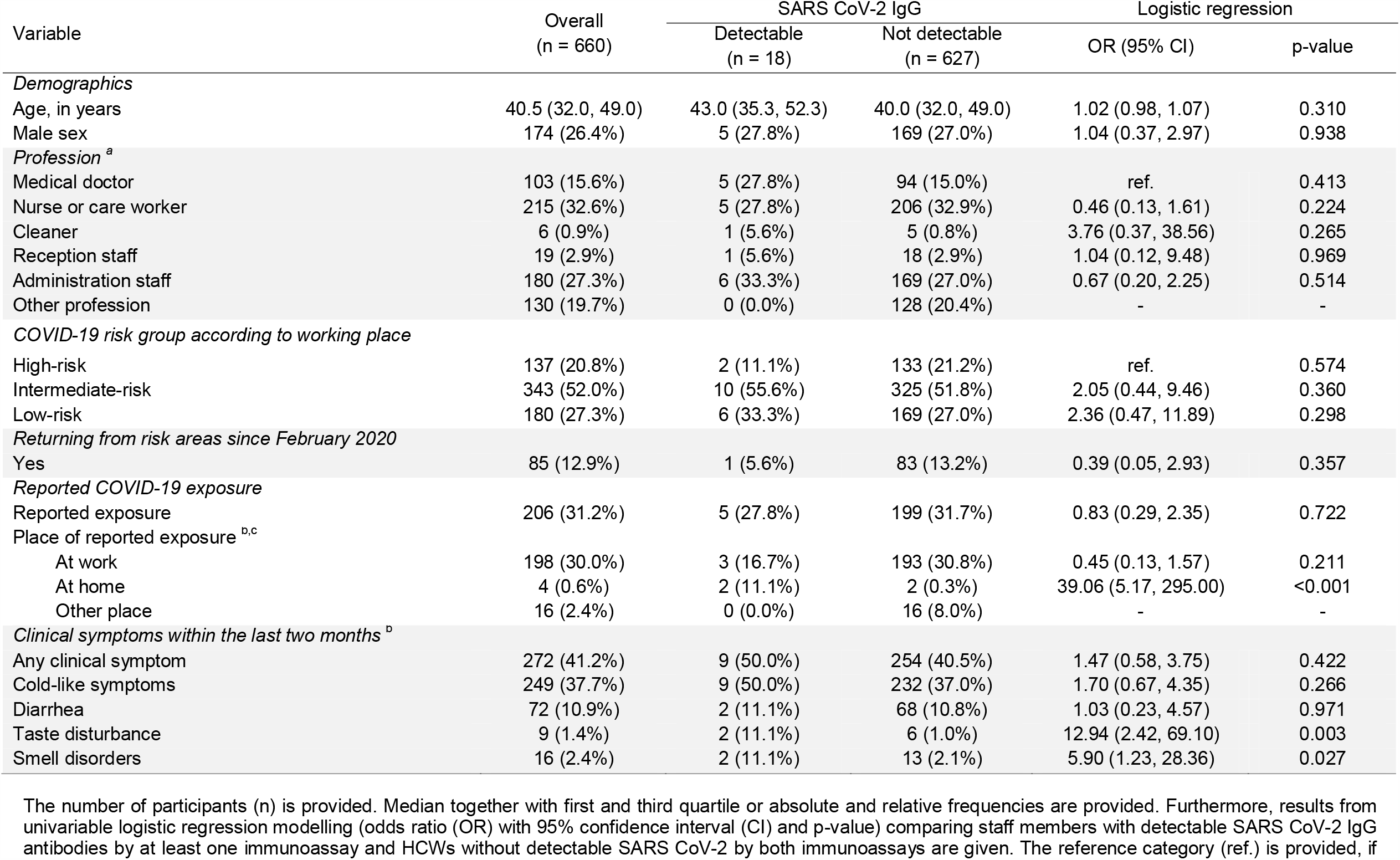
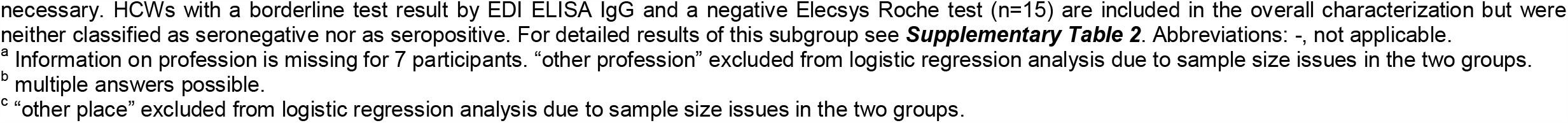
Characteristics of the study population – overall and stratified by antibody test result (SARS CoV-2 IgG)

| Variable | Overall<br>(n = 660) | SARS CoV-2 IgG |  | Logistic regression |  |
| --- | --- | --- | --- | --- | --- |
|  |  | Detectable<br>(n = 18) | Not detectable<br>(n = 627) | OR (95% CI) | p-value |
| *Demographics* |  |  |  |  |  |
| Age, in years | 40.5 (32.0, 49.0) | 43.0 (35.3, 52.3) | 40.0 (32.0, 49.0) | 1.02 (0.98, 1.07) | 0.310 |
| Male sex | 174 (26.4%) | 5 (27.8%) | 169 (27.0%) | 1.04 (0.37, 2.97) | 0.938 |
| *Profession <sup>a</sup>* |  |  |  |  |  |
| Medical doctor | 103 (15.6%) | 5 (27.8%) | 94 (15.0%) | ref. | 0.413 |
| Nurse or care worker | 215 (32.6%) | 5 (27.8%) | 206 (32.9%) | 0.46 (0.13, 1.61) | 0.224 |
| Cleaner | 6 (0.9%) | 1 (5.6%) | 5 (0.8%) | 3.76 (0.37, 38.56) | 0.265 |
| Reception staff | 19 (2.9%) | 1 (5.6%) | 18 (2.9%) | 1.04 (0.12, 9.48) | 0.969 |
| Administration staff | 180 (27.3%) | 6 (33.3%) | 169 (27.0%) | 0.67 (0.20, 2.25) | 0.514 |
| Other profession | 130 (19.7%) | 0 (0.0%) | 128 (20.4%) | - | - |
| *COVID-19 risk group according to working place* |  |  |  |  |  |
| High-risk | 137 (20.8%) | 2 (11.1%) | 133 (21.2%) | ref. | 0.574 |
| Intermediate-risk | 343 (52.0%) | 10 (55.6%) | 325 (51.8%) | 2.05 (0.44, 9.46) | 0.360 |
| Low-risk | 180 (27.3%) | 6 (33.3%) | 169 (27.0%) | 2.36 (0.47, 11.89) | 0.298 |
| *Returning from risk areas since February 2020* |  |  |  |  |  |
| Yes | 85 (12.9%) | 1 (5.6%) | 83 (13.2%) | 0.39 (0.05, 2.93) | 0.357 |
| *Reported COVID-19 exposure* |  |  |  |  |  |
| Reported exposure | 206 (31.2%) | 5 (27.8%) | 199 (31.7%) | 0.83 (0.29, 2.35) | 0.722 |
| *Place of reported exposure <sup>b,c</sup>* |  |  |  |  |  |
| At work | 198 (30.0%) | 3 (16.7%) | 193 (30.8%) | 0.45 (0.13, 1.57) | 0.211 |
| At home | 4 (0.6%) | 2 (11.1%) | 2 (0.3%) | 39.06 (5.17, 295.00) | <0.001 |
| Other place | 16 (2.4%) | 0 (0.0%) | 16 (8.0%) | - | - |
| *Clinical symptoms within the last two months <sup>d</sup>* |  |  |  |  |  |
| Any clinical symptom | 272 (41.2%) | 9 (50.0%) | 254 (40.5%) | 1.47 (0.58, 3.75) | 0.422 |
| Cold-like symptoms | 249 (37.7%) | 9 (50.0%) | 232 (37.0%) | 1.70 (0.67, 4.35) | 0.266 |
| Diarrhea | 72 (10.9%) | 2 (11.1%) | 68 (10.8%) | 1.03 (0.23, 4.57) | 0.971 |
| Taste disturbance | 9 (1.4%) | 2 (11.1%) | 6 (1.0%) | 12.94 (2.42, 69.10) | 0.003 |
| Smell disorders | 16 (2.4%) | 2 (11.1%) | 13 (2.1%) | 5.90 (1.23, 28.36) | 0.027 |

The number of participants (n) is provided. Median together with first and third quartile or absolute and relative frequencies are provided. Furthermore, results from univariable logistic regression modelling (odds ratio (OR) with 95% confidence interval (CI) and p-value) comparing staff members with detectable SARS CoV-2 IgG antibodies by at least one immunoassay and HCWs without detectable SARS CoV-2 by both immunoassays are given. The reference category (ref.) is provided, if
<sup>a</sup> Information on profession is missing for 7 participants. "other profession" excluded from logistic regression analysis due to sample size issues in the two groups.
<sup>b</sup> multiple answers possible.
<sup>c</sup> "other place" excluded from logistic regression analysis due to sample size issues in the two groups.

Individuals working outside the precategorized risk areas (namely lab personal where at least a proportion deals with COVID-19 related clinical specimens but has no patient contact) or participating outside the planned study period were excluded.

After pseudonymization at the study center, blood samples were sent to the Department of Clinical Chemistry and Laboratory Medicine (JUH) and the Institute of Medical Microbiology (JUH) for testing of IgG antibodies against SARS-CoV-2 by two different immunoassays (see below).

Pseudonymized questionnaires were digitalized with support of data management of the Institute of General Practice and Family Medicine (JUH). After digitalization, the whole data set was checked for plausibility and obvious errors were corrected.

### Questionnaire

The questionnaire included questions on demographics, working area, individual exposure to confirmed COVID-19 cases, return from COVID-19 risk areas since February 2020, results of previous polymerase chain reaction (PCR) or serology test for COVID-19, clinical symptoms within the last two months such as cold-like symptoms, diarrhea, taste disturbances and smell disorders. The severity of respiratory symptoms, included in the Wisconsin Upper Respiratory Symptom Survey (WURSS-24), was asked. WURSS-24 is identical to the WURSS-21 [22], except for the addition of the items assessing headache, body ache and fever.

To evaluate the risk for nosocomial transmissions, HCWs with an individual face-to-face contact within 1 meter with a confirmed COVID-19 patient or at least with its surroundings received an extended questionnaire that also included questions on the compliance concerning use of personal protective equipment (PPE) as recently published by the WHO (https://www.who.int/publications/i/item/risk-assessment-and-management-of-exposure-of-health-care-workers-in-the-context-of-covid-19-interim-guidance).

### SARS-CoV-2 antibody testing

Presence of SARS CoV-2 antibodies was investigated by two different commercially available IgG detecting immunoassays: an enzyme-linked immunosorbent assay EDI Novel Coronavirus SARS-CoV-2 IgG ELISA (Epitope Diagnostics Inc., San Diego, USA) and a chemiluminescence-based immunoassay Elecsys Anti-SARS-CoV-2 (Roche, Basel, Switzerland). Both assays target recombinant nucleocapsid protein and were carried out according to the manufacturers’ instructions. Sensitivities and specificities as provided by the manufacturers are high for both tests (≥98%). In case of two corresponding negative test results by both immunoassays, the participant was regarded as SARS-CoV-2 seronegative. Volunteers with at least one positive test result were regarded as SARS-CoV-2 seropositive. In case of a “borderline” test result for EDI IgG ELISA and a negative Elecsys Roche test, the test persons was neither classified as SARS-CoV-2 seropositive nor seronegative. Re-testing was offered to all participants with a divergent result of both immunoassays.

### Outcomes and further definitions

The primary outcome of the study was to assess the seroprevalence of SARS-CoV-2 antibodies in employees using two IgG detecting immunoassays. Secondary outcomes were (i) seroprevalence rates stratified by their risk of COVID-19 exposure during work (see below for definition of COVID-19 risk areas), (ii) potential risk factors and clinical symptoms for seropositive employees and (iii) compliance of HCWs in high-risk and intermediate-risk areas to wear PPE in case of an individual reported contact with a confirmed COVID-19 positive patient or its surroundings.

We classified employees in three groups according to their risk of a contact with COVID-19 patients: low, intermediate and high risk. The low-risk group included employees working in the administration without patient contact. The intermediate-risk group had regular patient contact but did not routinely treat patients with confirmed or suspected SARS-CoV-2 infections. The high-risk group included HCWs working at areas with confirmed COVID-19 patients and areas that deal with a high number of suspected COVID-19 cases (see ***Supplementary Table 1***).

### Sample size considerations

As previous data on SARS CoV-2 IgG seroprevalence rates of HCWs in Germany were sparse [23], our intention was to conduct an exploratory study focussing on the precision of the prevalence estimate in the defined exposure groups (i.e. the group comparisons by hypothesis test was not the primary objective). Thus, we assumed a true prevalence of 5%. 150 employees per group should be targeted to get 95% confidence intervals (for the proportion) with a precision (half width of confidence interval) of about 3.5%.

### Statistical analysis

Characteristics of participants are summarized (overall, stratified by test result) as absolute and relative frequencies or as median together with first and third quartile (Q1, Q3). Seroprevalence of SARS-CoV-2 in employees are described with absolute and relative frequencies together with 95% Clopper-Pearson confidence intervals (CIs). To compare seroprevalence rates between participants working at different COVID-19 risk areas, to analyze clinical symptoms and to identify potential risk factors for seropositive compared to seronegative participants, we apply univariable logistic regression modelling with the seropositivity as dependent variable and the investigated factor as independent variable. We provide odds ratios (OR) together with 95% CI. Compliance of HCWs (in high and intermediate-risk areas) to wear PPE is assessed with Fisher exact test. We compare those HCWs who stated to always or mostly wear PPE to those who stated not to wear PPE or did not provide information on this issue.

We applied a two-sided significance level of 0.05 and did not correct for multiple testing as all analyses were considered exploratory. Clopper-Pearson CIs were calculated with Microsoft Excel 2016. All other analyses were done with SPSS Statistics version 25.0 for Windows (IBM Corp., Armonk, NY, USA).

## Results

### Characteristics of the study population

We identified 3,228 employees who were eligible for study inclusion. Among them, 721 participants (22.3%) were included, and 660 of 721 participants (91.5%) could be analysed (see Figure 1). Of the 660 analysed participants, 174 (26.4%) were males and 486 (73.6%) were females. The median age of the participants was 40.5 (Q1-Q3: 32.0-49.0) years. The most common professions involved included nurses (n=215, 32.6%), followed by administration staff (n=180, 27.3%) and medical doctors (n=103, 15.6%), nursing assistants (n=18; 2.7%), psychologists (n=18; 2.7%) and ergo therapists (n=17; 2.6%). Two-hundred six employees (31.2%) reported direct exposure to a confirmed COVID-19 case. Among 198 employees with direct COVID-19 contact at work, 12 participants (6.1%) reported contact to a SARS-CoV-2 positive colleague. Direct COVID-19 contact outside the JUH included household contacts (n=4), patient contacts at other health care facilities (n=6), in the ambulance (n=5) or at a home visit (n=1), direct contact in town (n=2), at vacation (n=1) or with a former employee (n=1). Further details on the participants are provided in ***Table 1***.

**Figure 1:**
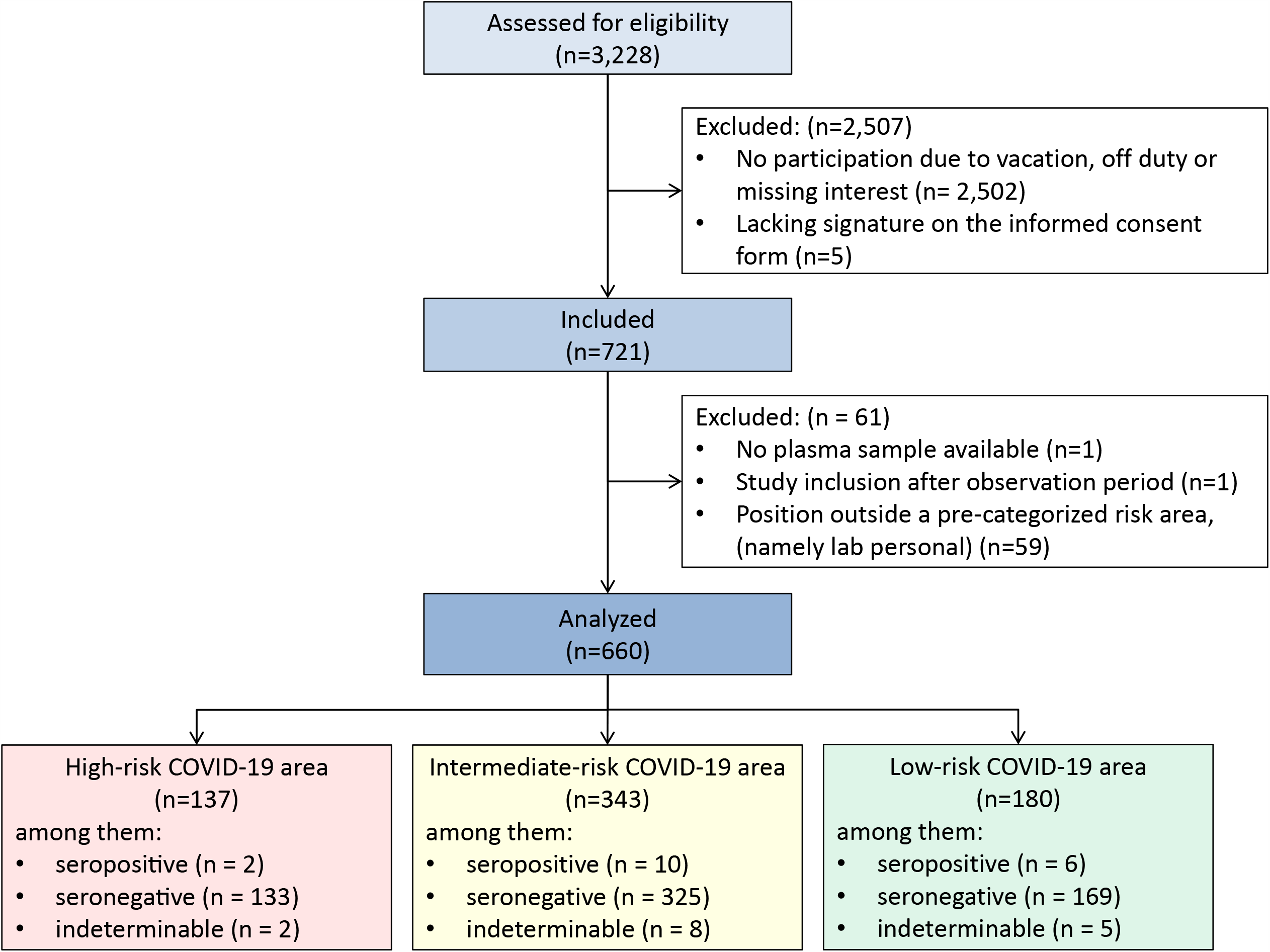
Flow chart of the Co-HCW study. The number of employees (n) are provided. Reasons for exclusions are given. Employees working at predefined areas at Jena University Hospital (JUH) were eligible for study inclusion. Working areas were classified into three categories according to the risk to deal with COVID-19 positive patients (see Supplementary Table 1 for the definition). Note that we decided not to assign lab personal to a pre-categorized risk area because a proportion dealt with COVID-19 related clinical specimens but there was no patient contact.

### Seroprevalence, infection status and previous testing for COVID-19

Among the 660 participants, 627 (95.0%) were tested negative for SARS-CoV-2 IgG antibodies by both immunoassays. Two participants (0.3%) were tested positive by Elecsys Roche and EDI IgG ELISA and 16 participants (2.4%) were only tested positive by EDI IgG ELISA. Fifteen participants had a “borderline” result for EDI IgG ELISA. Hence, 18 employees (2.7%, 95% CI 1.6%-4.3%), 12 HCWs (2.5% within the group HCWs, 95% CI 1.3%-4.3%) and 6 administration employees (3.3% within the group administration employees, 95% CI 1.2%-7.1%) had detectable SARS-CoV-2 IgG antibodies in at least one immunoassay. When considering also previously reported PCR and serology results, cumulative SARS-CoV-2 infection rate in our participants was 3.2% (95% CI 2.0%-4.8%).

Among the 18 employees with detectable SARS-CoV-2 IgG antibodies, only 9 (50.0%) reported clinical symptoms within the last two months (see ***Table 2***).

**Table 2.** Current and reported test results and clinical symptoms for COVID-19 in employees of Jena University Hospital stratified by (a) detectable antibodies after recruitment, (b) history of past positive SARS-CoV-2 polymerase chain reaction (PCR) or serology, and (c) any evidence of a COVID-19 infection

|  | Employees seropositive for<br>SARS-CoV-2 IgG antibodies<br>after recruitment | Employees with reported evidence<br>of a positive SARS-CoV-2 test<br>prior recruitment | Employees with any evidence of<br>past/current SARS-CoV-2 infection |
| --- | --- | --- | --- |
| *Overall* |  |  |  |
| Number of Employees | 18 out of 660 tested | 4 out of 212 tested | 21 out of 660 tested |
| Proportion (95% CI) | 2.7% (1.6% to 4.3%) | 1.9% (0.5% to 4.8%) | 3.2% (2.0% to 4.8%) |
| *Among respective Employees* |  |  |  |
| Not previously diagnosed as COVID-19 by<br>PCR or serology | 17 (94.4%, 72.7% to 99.9%) | - | 17 (81.0%, 58.1% to 94.6%) |
| COVID-19 symptoms reported | 9 (50.0%, 26.0% to 74.0%) | 4 (100%, 38.8% to 100.0%) | 12 (57.1%, 34.0% to 78.2%) |
| Maximum severity of cold-like symptoms<br>within the last two months | 0.5 (0.0, 4.0) | 3.0 (1.3, 4.0) | 1.0 (0.0, 4.0) |

Absolute and relative frequencies together with 95% Clopper-Pearson confidence intervals (CI) or median together with first and third quartile are reported. Severity of illness (cold-like symptoms) is defined according to the Wisconsin Upper Respiratory Symptom Survey (0=no illness, 1-2=very mild, 3-4=mild, 5-6=moderate, 7=severe). Abbreviations: -, not applicable.

### Follow-up of discrepant immunoassay results

Thirteen of 15 participants (86.7%) with a “borderline” test result for EDI IgG ELISA and 12 of 16 participants (75.0%) with a positive result for EDI IgG ELISA but negative Elecsys Roche test were retested after 1-2 months. All employees with a previous “borderline” test result for EDI IgG ELISA became negative in the retest. For clinical symptoms in this group, we refer to ***Supplementary Table 2***. Three of 12 employees (25.0%) with a previous positive test result for EDI IgG ELISA but negative Elecsys Roche test had unchanged test results 1-2 months later, whereas 9 of 12 employees (75.0%) became negative in the retest. All 3 participants (2 HCWs and 1 administration employee) with a persistent positive test result for EDI IgG ELISA but negative Elecsys Roche test did not report any clinical symptoms.

### Potential risk factors and clinical symptoms for antibody positivity of employees

As shown in ***Table 1***, we did not observe evidence for an association of antibody positivity with the demographics, the professions or COVID-19 risk area (all p-values from logistic regression >0.05). The two persons who were tested positive by both immunoassays were administration employees. The only parameters that were associated with SARS-CoV-2 seropositivity in employees included close COVID-19 contact at home (OR 39.06, 95% CI 5.17 to 295.00), taste disturbances (OR 12.94, 95% CI 2.42 to 69.10) and smell disorders (OR 5.90, 95% CI 1.23 to 28.36).

### Compliance to wear PPE in case of a individual contact with a COVID-19 positive patient and/or its surroundings

Reported compliance to wear PPE was associated with the COVID-19 risk area according to the working place (p<0.001). Compliance of HCWs working in COVID-19 high-risk was 98.3% (yes: n=114; no: n=2) and in intermediate-risk areas 69.8% (yes: n=62, most time: n=5, no: n=21, no answer given: n=8). Detailed information on compliance to wear different items of PPE and different circumstances is given in ***Figure 2***.

**Figure 2.**
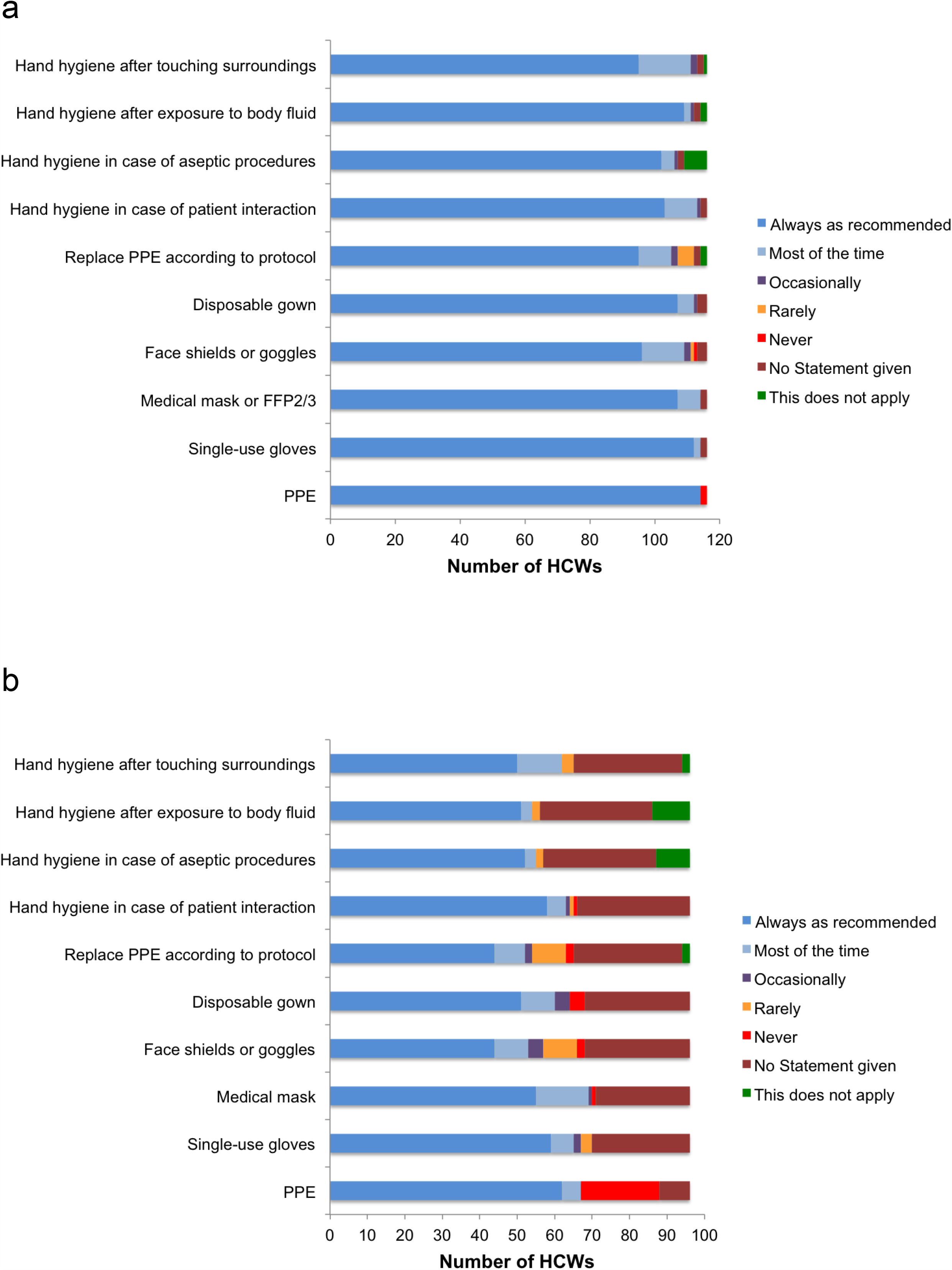
Compliance to wear personal protective equipment (PPE) in case of an individual reported contact with a confirmed COVID-19 positive patient or its surroundings in healthcare workers (HCWs) from (a) high-risk COVID-19 areas (n=116 HCWs) versus (b) intermediate-risk COVID-19 areas (n=96 HCWs). The definitions of risk areas are provided in ***Supplementary Table 1***.

A single HCW reported a puncture/sharp accident with material contaminated with biological fluid/respiratory secretions during a healthcare interaction with a COVID-19 patient. This HCW remained seronegative.

## Discussion

Altogether, seroprevalence of SARS-CoV-2 IgG was low in the investigated group (2.7%). Even when we also considered reported previous PCR and serology results, the cumulative SARS-CoV-2 infection rate remained below the expected 5% derived from reports of a similar University hospital in Münster, Germany [23]. Higher seroprevalence rates among frontline healthcare personnel were reported by recent studies outside Germany (ranging from 4.0% to 9.0%) [24–26].

Among 660 tested employees of JUH, we did not detect an association between seroprevalence and risk area according to their workplace. Contrary, HCWs working at COVID-19 high-risk areas had the numerically lowest (1.5%) and employees without any patient contact had the numerically highest seroprevalence rates (3.3%).

We found an association between seroprevalence rates and a reported individual exposure to a SARS-CoV-2 positive household contact. Similarly, Luo *et al*. who evaluated the risk for transmission in 3,410 close contacts of COVID-19 patients identified household contact as the main setting for transmission of SARS-CoV-2 (10.3%) [27]. Community acquisition is hence a major aspect that needs to be considered.

In the present study, HCWs in the high-risk group reported a remarkable compliance of 98.2% regarding PPE administration whereas compliance in the intermediate-risk group was significantly lower with 69.8%. Consequent use of PPE is an effective measure to minimise the risk of infection [20, 24]. We assume that the awareness regarding personal protection was higher in those HCWs who are repeatedly exposed to COVID-19 patients. An increased awareness might lead to better adherence to other hygienic measurements as well [28]. In addition, a daily routine in PPE use improves correct donning and doffing and thus, reduces the risk of contamination. The results of the study support the importance of adequate PPE use to prevent transmission from patient to HCW. Additionally, mandatory masking might have reduced nosocomial transmissions also in employees without patient contact. However, compliance to mandatory masking during working hours was not evaluated in our study. According to Wang *et al*., implementation of mandatory masking of HCWs and patients can be effective to reduce SARS-CoV-2 infection rates [29]. In a recently published report by Self *et al*. that included 3,248 frontline HCWs, seroprevalence of SARS-CoV-2 antibodies was lower among HCWs who reported always wearing a face covering while caring for patients compared to those who did not (6% versus 9%) [24].

It is known, that pre- and asymptomatic COVID-19 infected persons can be contagious despite absence of any subjective feeling of illness. In a population-based study by Gudbjartsson *et al*. including 30,576 people from Iceland, nearly one third of the SARS-CoV-2 infections were asymptomatic and durability of SARS-CoV-2 antibody levels was over 4 months [30]. In our study population, only 50% of those tested positive reported any clinical symptoms, but the presence of taste disturbances or smell disorders were both associated with seropositivity of SARS-CoV-2 IgG. Similarly, the authors Iversen *et al*. identified loss of smell or taste as the symptom that was most strongly associated with seropositivity in HCWs in Denmark [26]. In a complete cohort study following up a COVID-19 outbreak in a German village, we found that loss of taste and smell were the symptoms exhibiting the highest association with PCR positivity [21].

The validity regarding sensitivity and specificity of SARS-CoV-2 serology testing has not yet been investigated entirely [31]. According to the recently published IDSA guidelines on the diagnosis of COVID-19 there will be false positive and false negative tests, but the most reliable spot of measuring SARS-CoV-2 antibodies is 3-4 weeks after exposure to the virus/onset of clinical symptoms [32]. In the present study we used two different immunoassays, the Elecsys Anti-SARS CoV-2 Roche Diagnostics and the EDI Novel Coronavirus SARS-CoV-2 IgG ELISA. A recent head-to-head comparison of both immunoassays found acceptable agreement between both tests [33]. In our study the Roche Elecsys assay did not identify asymptomatic COVID-19 cases, which was already observed in the CoNAN study mentioned above [21]. This assay was only positive in 2 participants (0.3%) who were tested at beginning of June 2020 and had developed a symptomatic COVID-19 disease in the second half of March 2020. In contrast, the EDI IgG ELISA was positive in 2.7% of participants (9 asymptomatic and 9 symptomatic cases) including the two persons with the positive Roche Elecsys assay. It is still a matter of debate to what extend seroprevalence of SARS-CoV-2 IgG can really reflect immunity and status after infection. Nonetheless, serological examination is a method which is easily available and highly cost-effective compared to PCR [34].

This study has the following limitations: Despite the high number of participants, the recruitment rate was below 10% of the total JUH staff and results of previous COVID-19 testing and compliance using PPE were only recorded by self-reports. We determined antibody titres repeatedly only in those with discrepant results. As SARS-CoV-2 infection generates two waves of antibodies, the provided data do not reflect long-lived immunity [34].

## Conclusion

In our study, reported contact with a COVID-19 patient was not found to be a risk factor for seroprevalence of SARS CoV-2 antibodies, whereas contacts with infected family members was highly predictive. In line, we found a high awareness and compliance with PPE and no evidence for higher seroprevalence in HCWs caring for COVID-19 patients, whereas administration employees with no patient contacts had numerically higher seroprevalence rates. We conclude that for HCWs, community transmission may play a larger role for COVID-19 infection than professional exposure when using appropriate PPE.

## Data Availability

The data that support the findings of this study are available from the authors upon reasonable request.

## Acknowledgements

We thank Stefanie Baier, Jana Schmidt, Stefanie Kolanos and Monique Philippe for excellent technical support.

## Author contributions

CB, AK, MWP had full access to all of the data in the study and take responsibility for the integrity of the data and the accuracy of the data analysis.

Study concept and design: SW, MWP, ASch, JM, BL, MKi, ASta, ASte, MBau, WB, FZ, MW. Acquisition of data: CB, AK, SH, JA.

Performing of seroprevalence testing: CR, MKi, MBai, BL.

Statistical analyses: CB, MKe.

Drafting of the manuscript: CB, AK, MKe, MWP.

Critical revision of the manuscript and additional important intellectual content, data interpretation: all authors.

Study supervision: MWP, CB.

